# Energy Landscape and Dynamic State Modeling for Schizophrenia Biomarker Extraction in Task-Based fMRI

**DOI:** 10.64898/2025.12.14.25342238

**Authors:** Janerra D. Allen, L. Elliot Hong, Fow-Sen Choa

## Abstract

Task-based functional magnetic resonance imaging (fMRI) examines how the brain dynamically responds to cognitive and perceptual demands, offering complementary insight beyond traditional activation-based analyses in schizophrenia. Prior task-based fMRI studies have identified reduced functional connectivity within auditory and associated cortical areas. In this study, we investigated task-evoked functional connectivity and brain state dynamics in 25 healthy controls, 23 patients with schizophrenia experiencing auditory verbal hallucinations (AVH+), and 23 patients without hallucinations (AVH-). Participants completed multiple auditory paradigms, including word lists, sentence lists, and reverse speech. The most robust connectivity differences emerged during the word list task, where cluster-level analyses revealed distinct network interaction patterns differentiating controls, AVH+, and AVH-patients. Energy landscape modeling further demonstrated altered stability and organization of brain states in both patient groups, with the largest deviations observed in AVH+ individuals. These alterations point to dysregulated neural dynamics linked to hallucination vulnerability. Overall, the results show that task-based fMRI sensitively captures disruptions in functional connectivity and brain state stability relevant to auditory hallucinations, underscoring its value for characterizing network-level mechanisms underlying perceptual disturbances in schizophrenia.

## Introduction

Functional magnetic resonance imaging (fMRI) has become a cornerstone of human brain mapping, providing critical insights into intrinsic and stimulus-driven neural processes. Resting-state fMRI (rs-fMRI) has revealed consistent patterns of functional connectivity and large-scale brain networks, offering a foundational view of neural organization in both healthy and diseased brains [1, 2]. However, resting-state paradigms lack explicit task demands and are limited in capturing dynamic, context-dependent neural responses that underpin cognition and psychiatric symptoms. Moreover, dynamic functional connectivity analyses have revealed that resting-state networks exhibit time-varying patterns, underscoring the need for task-based paradigms to capture transient cognitive states [3]. In contrast, task-based fMRI (tb-fMRI) enables researchers to probe how specific cognitive, perceptual, or motor tasks engage distributed brain networks, resulting in behaviorally anchored neural signatures [4, 5].

Schizophrenia, a complex neuropsychiatric disorder marked by disturbances in perception, cognition, and behavior, presents a compelling case for task-based neuroimaging. Although rs-fMRI studies have highlighted widespread dysconnectivity in default mode, sensorimotor, and thalamocortical networks, these findings often provide generalized signatures that lack specificity for individual symptoms or subtypes [6, 7]. Task-based approaches help address this gap by capturing how brain networks respond to cognitive load, offering more precise insights into the neural mechanisms underlying core symptoms such as auditory verbal hallucinations (AVH), delusions, and working memory impairments. Recent studies using symptom-capture and neurofeedback paradigms have demonstrated that task-based fMRI can isolate AVH-specific network disruptions, particularly in frontotemporal and cingulate regions [8–10].

Among these, AVH remains one of the most intriguing and debilitating symptoms. Hallucinations reflect a failure in the brain’s ability to distinguish internally generated representations, such as inner speech, from externally driven sensory input.

Neuroimaging studies have consistently implicated abnormal activation in the auditory cortex, speech-motor areas, and prefrontal regions during AVH episodes [11]. This supports the view that hallucinations arise from dysfunctions in self-monitoring, sensory integration, and internal model updating. These dysfunctions are consistent with predictive coding frameworks, which posit that hallucinations are the result of an imbalance between top-down priors and bottom-up sensory input [12, 13].

Crucially, hallucinations are not isolated errors in perception, but dynamic processes shaped by brain state transitions. They may arise when unstable or aberrant network dynamics leads to spontaneous activations of sensory representations, even in the absence of corresponding stimuli. From a systems-level perspective, these dynamics result from impaired functional connectivity, particularly between control hubs (e.g., prefrontal cortex) and sensory areas—combined with a failure to suppress internally generated predictions. Task-based fMRI is uniquely positioned to reveal these mechanisms by forcing the brain to engage with structured stimuli and observing deviations from normative responses in real time.

To characterize these dynamics, this study applies energy landscape modeling, a framework drawn from statistical physics that visualizes brain activity as traversals through a landscape of stable and unstable states. In this model, brain states correspond to attractors in a high-dimensional space, and transitions reflect shifts in network configurations. In healthy brains, tasks evoke transitions into stable, goal-directed states. In schizophrenia, especially in individuals with AVH, these landscapes may become shallow, noisy, or fragmented, making the brain more susceptible to falling into maladaptive or hallucinatory attractors. Previous applications of energy landscape analysis in schizophrenia have identified fragmented attractor states and reduced network stability, supporting its utility in modeling psychiatric symptoms [14, 15].

This study takes advantage of these parallels to investigate hallucination generation from a unified perspective. We analyze task-evoked fMRI data from three groups: healthy controls, schizophrenia patients with AVH (AVH+) and those without AVH (AVH-). Using language-processing tasks known to activate the auditory and prefrontal regions implicated in AVH, we examine how brain activation, functional connectivity, and dynamic transitions differ between groups.

Our approach integrates task-based fMRI with energy landscape modeling to characterize alterations in the stability and variability of functional brain states. By comparing hallucinating and non-hallucinating patients, we aim to identify neural signatures that distinguish pathological internal representation from normative perceptual processing.

This framework provides a novel account of AVH as a failure of context-sensitive generative control. Beyond deepening our understanding of schizophrenia, the results may contribute to a broader theory of hallucination as a failure mode in predictive systems—biological or artificial.

## Materials and methods

### Data Characteristics

Twenty-five healthy controls and forty-six individuals diagnosed with schizophrenia were included in this study, as detailed in Table 1. The data were obtained from the publicly available dataset associated with Soler-Vidal et al. (2022) [16]. The schizophrenia cohort was subdivided into two groups: 23 patients who frequently experienced auditory verbal hallucinations (AVH+) and 23 who did not report hallucinations (AVH-).

**Table 1.** Demographics of healthy controls and patients.

|  | Control vs Patient |  |  | AVH vs AVH+ |  |  |
| --- | --- | --- | --- | --- | --- | --- |
|  | CON<br>*N* = 25 | PAT<br>*N* = 46 | Differences<br>CON vs PAT | AVH-<br>*N* = 23 | AVH+<br>*N* = 23 | Differences<br>AVH- vs AVH+ |
| Age | 39.80 ± 14.08 | 42.52 ± 10.72 | *t* = -0.84, *p* = 0.40 | 44.96 ± 7.38 | 40.09 ± 12.96 | *t* = 1.57, *p* = 0.13 |
| Sex | 18 (Male)<br>7 (Female) | 36 (Male)<br>10 (Female) | $\chi^2$ = 0.35, *p* = 0.55 | 16 (Male)<br>7 (Female) | 20 (Male)<br>3 (Female) | $\chi^2$ = 2.04, *p* = 0.15 |
| IQ | 101.17 ± 9.91 | 99.82 ± 9.27 | *t* = 0.55, *p* = 0.59 | 101.32 ± 8.89 | 98.39 ± 9.60 | *t* = 1.06, *p* = 0.29 |
| PSYRAT-H | n/a | 12.46 ± 13.60 | *t* = n/a, *p* = n/a | 0 | 24.91 ± 7.32 | *t* = -16.32, *p* < 0 |

IQ: intelligence quotient; PSYRATS-H: Psychotic Symptom Rating Scale for hallucination severity. Values are means (± SD) unless otherwise noted.

### Data Acquisition

The original participants were recruited from psychiatric services in Spain, as reported by Soler-Vidal et al. The study received ethical approval from the Research Ethics Committee FIDMAG Sisters Hospitallers. Our analysis used fully de-identified, publicly available data; the authors of this study had no access to information that could identify individual participants. The dataset was accessed for research purposes in approximately May 2024.

The dataset includes functional MRI data collected during a block design consisting of three auditory experimental conditions: word lists, sentence lists, and reverse speech. Clinical measures included estimated IQ and the Psychotic Symptom Rating Scale for Auditory Hallucinations (PSYRATS-H), a validated multidimensional scale assessing AVH features such as frequency, distress, and controllability [17, 18].

Although all three tasks were collected and initially analyzed, this study focuses primarily on the word list task. This choice was made a priori because the word list paradigm provides the strongest and most consistent engagement of auditory and language networks implicated in AVH, including primary auditory cortex, superior temporal gyrus, and prefrontal monitoring regions. These regions are central to prevailing mechanistic models of auditory hallucinations and to the hypotheses of this study. The focus on this task therefore reflects theoretical relevance and neural specificity, not data-driven selection.

### Data Preprocessing and Denoising

Preprocessing followed the approach described by Allen et al. [19], using the CONN and SPM toolboxes for functional connectivity analysis, as detailed in the CONN handbook [20, 21]. Steps included anatomical CompCor denoising, task regressors, and ROI-to-ROI clustering methods tailored for task-based designs. Adjustments were made to incorporate stimulus-event regressors and to model condition-specific BOLD dynamics. The CONN toolbox was used for both block design preprocessing and structured denoising, isolating task-related neural fluctuations from intrinsic activity. Denoising incorporated anatomical CompCor, a component-based noise correction method that reduces physiological and motion-related artifacts [22].

### First-Level Analysis

At the individual level, functional connectivity was computed using 246 regions of interest (ROIs) from the Brainnetome atlas [23]. The seed-to-voxel and ROI-to-ROI connectivity metrics were calculated using Fisher-transformed bivariate correlation coefficients, modeled via a weighted general linear model (GLM) [21]. Task regressors were convolved with the canonical hemodynamic response function to account for condition-specific BOLD fluctuations. Condition-specific activation and connectivity were estimated using the GLM framework [24], with scan weights adjusted for task timing to capture transient neural responses.

### Second-Level (Group-Level) Analysis

Group-level analysis used a GLM to compare task-modulated connectivity across groups. Functional Network Connectivity (FNC) and Spatial Pairwise Connectivity (SPC) were used to define and analyze connection clusters, adapted from the methods used in resting-state studies [25, 26]. Connectivity estimates were derived from contrast maps that isolate condition-specific responses to auditory stimuli. Statistical inference was performed using Random Field Theory to control for family-wise error rates [26].

#### Functional Network Connectivity

Functional network connectivity (FNC) was used to analyze groups of functionally related ROIs, using hierarchical clustering to define connectivity networks based on anatomical proximity and task-dependent functional similarity. Connectivity matrices were examined for patterns within and between networks, revealing task-modulated connectivity effects. FNC statistics included F-values for each ROI pair, with uncorrected and FDR-corrected p-values to identify significant clusters. Task-related changes in functional connectivity were visualized as connectome rings, highlighting significant ROI-to-ROI correlations [27].

#### Spatial Pairwise Connectivity

Spatial pairwise connectivity (SPC) was applied to the ROI-to-ROI connectivity matrices for each task condition, using optimal leaf ordering to refine hierarchical clustering and improve cluster interpretability [28]. Analysis followed the same cluster-wise inference framework as the resting-state literature, with connectivity differences assessed based on task conditions. SPC results included T- or F-statistic maps thresholded to identify significant clusters, with uncorrected and FDR-corrected p-values capturing task-modulated connectivity patterns. Suprathreshold clusters were iteratively refined to estimate the probability density of task-related connectivity effects under the null hypothesis, ensuring robust detection of stimulus-driven functional connectivity changes [27]. Cluster significance was assessed using topological false discovery rate (FDR) correction, which accounts for spatial dependencies in neuroimaging data [27].

In CONN’s SPC clustering inference measure, the terms score, mass, and size describe different properties of connectivity clusters. The score reflects the strength of individual connections and is typically represented as a t-score or z-value. The mass summarizes the total accumulated connectivity strength across the cluster, and the the size reflects to the number of connections within the cluster. Thus, score indicates connection strength, size reflects spatial extent, and mass provides an integrated measure of both.

Together, these statistical frameworks—GLM modeling, Random Field Theory, topological FDR, and hierarchical clustering—ensure robust detection of task-driven connectivity differences across clinical groups.

### Statistical Testing Procedures

To assess task-dependent functional connectivity differences across groups, second-level analyses were performed using general linear models (GLMs). Connectivity estimates were derived from contrast maps isolating condition-specific responses. Two methods were used to identify significant clusters of ROI-to-ROI connectivity: Functional Network Connectivity (FNC) and Spatial Pairwise Connectivity (SPC), both implemented in the CONN toolbox.

Cluster-level inference was performed using F-statistics or T-statistics (depending on the comparison), followed by Bonferroni correction to account for multiple comparisons. For each task condition, the total number of possible clusters was used to adjust the significance threshold (e.g., *α* = 0.05 / total clusters), controlling for Type I error in the context of multiple testing.

### REX: Neuroimaging Toolbox

To prepare inputs for energy landscape modeling, regions of interest (ROIs) corresponding to Bonferroni-corrected significant clusters were extracted from the connectivity analyses. Task-based fMRI time series were obtained for each subject by spatially aligning the 246-region Brainnetome atlas [23] to denoised, post-processed functional images using REX, a MATLAB-based toolbox for ROI analysis [29]. This procedure yielded a matrix of dimensions 341 × 246, representing time points and ROI-specific BOLD signals. For subsequent analyses, we focused on a subset of ROIs identified through SPC-based clustering, which showed the strongest differences between groups following Bonferroni correction. The original matrix was transposed and subset by ROI and task condition to generate reduced time series matrices for each comparison group. These reduced matrices served as input to the energy landscape (EL) analysis.

### EL analysis

The energy landscape (EL) analysis toolbox was used to interpret the matrices of reduced ROIs, identified by Bonferroni-corrected clustering, along with their corresponding time series. Energy landscape analysis characterizes the possible states of a system, illustrating how configurations, such as neural activity patterns, evolve over time. This approach is based on the pairwise maximum entropy model (MEM), which fits second-order statistical models to empirical data to capture dynamic fluctuations in brain activity [14, 15, 30–33].

At each ROI, indexed by *N* , and at each time point, the fMRI signal was binarized into active or inactive states by thresholding the signal relative to the mean of each ROI.

The normalized frequency of each binary activity pattern was then computed. For *N* ROIs, there are 2*^N^* possible brain states. Using MEM, energy values and probabilities were estimated for each state. Energy and probability are inversely related: states with lower energy and higher probability occur more frequently and correspond to more stable brain configurations. The pairwise MEM is preferred for its balance between computational tractability and its ability to capture essential pairwise interactions while excluding higher-order dependencies.

Conceptually, the energy landscape provides a framework for visualizing the stability and transitions between brain states, where low-energy basins correspond to stable functional configurations, and transitions between basins reflect dynamic changes in connectivity. This framework enables the characterization of dynamic brain-state transitions and has been successfully applied in both resting-state and task-related studies [15, 30].

In the present analysis, EL modeling was applied to reduced ROI sets derived from both FNC and SPC clustering methods. This dimensionality reduction facilitated more interpretable state-space modeling while preserving key functional distinctions relevant to the differences between the task and symptom groups.

Comparisons based on FNC-derived ROIs included:

1. Controls vs. AVH+ patients during the sentence task (N=9)
2. Controls vs. AVH+ patients during the word task (N=9)
3. AVH+ vs. AVH- patients during the reverse speech task (N=5)
4. AVH+ vs. AVH- patients during the sentence task (N=7)
5. AVH+ vs. AVH- patients during the white noise condition (N=2)

Comparisons based on SPC-derived ROIs included:

1. Controls vs. AVH+ patients during the word task (N=5)
2. AVH+ vs. AVH- patients during the sentence task (N=6)
3. AVH+ vs. AVH- patients during the word task (N=6)

### Paired States

Each subject had 2*^N^* possible connectivity states, each associated with a specific energy and probability value. The energy values for all states were computed across all subjects. Two-sample *t*-tests were conducted to compare energy values between the control and AVH+ groups, as well as between the AVH+ and AVH- groups. Bonferroni correction was applied with a significance level of *α* = 0.05, adjusted for the number of possible states (2*^N^* ).

Complementary paired states were identified within each subgroup based on their energy profiles. These paired states represent distinct connectivity patterns and provide information on functional brain dynamics between conditions and participants.

### Energy-Based State Modeling

Task-based fMRI captures neural activity in response to cognitive demands, enabling the study of network-level dynamics. To characterize these dynamics, we applied energy landscape (EL) modeling, which represents patterns of brain activity as trajectories through a high-dimensional landscape of stable and unstable states, or attractors [14, 15, 30–33]. Low-energy states correspond to frequently visited, stable configurations, whereas high-energy states are transient and less frequently occupied.

EL modeling is grounded in the pairwise maximum entropy model (MEM), which has been validated in resting-state fMRI for capturing functional interactions and identifying reproducible attractor states [31, 32]. Bayesian estimation methods further allow individualized characterization of energy landscapes, providing insight into subject-specific variability in brain state dynamics [33].

While prior studies have primarily applied EL modeling to resting-state data, the relationship between energy landscape metrics—such as dwell times and transition probabilities—and cognitive or clinical features during task performance remains largely unexplored. Schizophrenia, particularly in patients with auditory verbal hallucinations (AVH+), is associated with deficits in attention, working memory, and sensory integration [3, 5]. These cognitive impairments may manifest as alterations in the stability and dynamics of functional brain states.

In this study, EL modeling was applied to task-based fMRI during the auditory word list task, focusing on regions identified through previous Functional Network Connectivity (FNC) and Spatial Pairwise Connectivity (SPC) analyses. We hypothesized that AVH+ patients would exhibit altered energy landscapes relative to controls and AVH- patients, reflected in differences in the stability (low-energy states), frequency of occurrence, and transition dynamics between functional states.

By quantifying these metrics, this approach provides a framework for exploring how cognitive processes—including attention, working memory, and sensory integration—may be disrupted in schizophrenia. Importantly, interpretations of energy-based states as reflecting cognitive stability or flexibility are exploratory and theoretically based rather than previously established clinical findings [14, 15, 30–33].

### State Transition Dynamics

Analyzing transitions between brain states provides insight into the dynamic reconfiguration of functional networks. Dwell time—the duration the system remains in a given brain state—is a commonly used metric in dynamic and energy landscape analyses and has been shown to vary between healthy and clinical populations [3, 14, 31–33]. Shorter dwell times may indicate rapid state shifts and network instability, which can contribute to cognitive deficits in psychiatric disorders.

Studies in schizophrenia suggest that AVH+ patients exhibit altered state dynamics, spending more time in hyperconnected or maladaptive states and less time in stable, goal-directed configuration [15, 30]. Energy-based metrics such as visit counts, dwell times, and transition probabilities quantify these differences and have been linked to behavior, cognition, and clinical outcomes [31, 32].

Based on this framework, we hypothesized that AVH+ individuals would exhibit distinct dwell-time distributions relative to AVH- and control groups, reflecting disrupted engagement of task-relevant brain states and impaired cognitive control.

## Results

### Group-Level Connectivity Results

We first examined ROI-to-ROI functional connectivity across all auditory tasks. Individual connectivity comparisons revealed no significant differences between controls and all patients, controls and AVH- patients, controls and AVH+, or between male and female participants. However, when considering average connectivity across clusters, significant differences emerged between controls and AVH+ patients, as well as between AVH- and AVH+ patients.

At the level of individual connections, no pairwise connectivity differences survived correction for multiple comparisons across groups (controls, AVH-, AVH+, male, female), consistent with previous observations that schizophrenia-related dysconnectivity is often distributed rather than localized [1, 3]. However, when considering average connectivity across clusters, significant differences emerged between controls and AVH+ participants, as well as between AVH- and AVH+ participants.

This pattern suggests that network-level shifts, rather than large isolated changes, underlie task-related dysconnectivity in schizophrenia.

Functional connectivity analyses were conducted across all auditory conditions, including word lists, sentence lists, reverse speech, and a white noise control.

Group-level comparisons were performed using both Functional Network Connectivity (FNC) and Spatial Pairwise Connectivity (SPC) approaches. While analyses were completed for all tasks, the word list task produced the most robust and statistically significant group differences. Accordingly, the following results focus primarily on findings from the word list task, with other task results noted only where relevant.

### Control vs. Patients

Comparing healthy controls (*N* = 25) to all patients with schizophrenia (*N* = 46), no significant clusters were observed in either FNC or SPC analyses across any task after Bonferroni correction.

### Control vs. AVH+ Patients

During the word list task, significant differences were detected. FNC identified 5 significant clusters comprising 11 ROIs, and SPC identified 1 cluster comprising 5 ROIs (see Tables 2 and 3). For the sentence list task, FNC identified 3 significant clusters and SPC identified 5, but these did not survive Bonferroni correction. No significant findings were observed for the reverse speech or white noise conditions.

**Table 2.** Statistical cluster analysis for the between-subject [Control (*−*1) vs. AVH+ (1)] contrast using functional network connectivity (FNC), presenting significant connectivity between nodes for the *word lists* task (post-Bonferroni correction).

| Analysis |  | Statistic | p-unc | p-FDR |
| --- | --- | --- | --- | --- |
| Cluster 1 |  | F(2,45) = 15.14 | < 0.00 | 0.01 |
| Left lPFtha | Right mPFtha | T(46) = 5.55 | < 0.00 | 0.00 |
| Right lPFtha | Right mPFtha | T(46) = 5.32 | < 0.00 | 0.00 |
| Right mPMtha | Right lPFtha | T(46) = 4.64 | < 0.00 | 0.01 |
| Cluster 2 |  | F(2,45) = 10.64 | < 0.00 | 0.05 |
| Left PPtha | Left V5/MT+ | T(46) = -4.77 | < 0.00 | 0.00 |
| Left PPtha | Right V5/MT+ | T(46) = -4.65 | < 0.00 | 0.00 |
| Cluster 3 |  | F(2,45) = 10.57 | < 0.00 | 0.05 |
| Right A4hf | Right A4t | T(46) = 4.62 | < 0.00 | 0.00 |
| Left A4hf | Left A4t | T(46) = 4.49 | < 0.00 | 0.01 |

p-unc: uncorrected p-value, p-FDR: false discovery rate-adjusted p-value.

**Table 3.** Statistical cluster analysis for the between-subject [CON (*−*1) vs. AVH+ (1)] contrast using spatial pairwise connectivity (SPC), presenting significant connectivity between nodes for the *word lists* task (post-Bonferroni correction).

| Analysis |  | Statistic | p-unc | p-FDR |
| --- | --- | --- | --- | --- |
| Cluster 1 |  | Score = 113.86 | 0.00 | 0.03 |
|  |  | Mass = 161.35 | 0.00 | 0.01 |
|  |  | Size = 6 | 0.00 | 0.03 |
| Left lPFtha | Right mPFtha | T(46) = 5.55 | < 0.00 | 0.03 |
| Right lPFtha | Right mPFtha | T(46) = 5.32 | < 0.00 | 0.03 |
| Right mPMtha | Right lPFtha | T(46) = 4.64 | < 0.00 | 0.09 |

p-unc: uncorrected p-value, p-FDR: false discovery rate-adjusted p-value.

### AVH+ vs. AVH- Patients

In this subgroup comparison (*N* = 23 each), FNC revealed significant clusters during the sentence (9/820) and reverse speech tasks (4/1035). SPC identified clusters during the sentence (9/1113) and word list tasks (5/1039). After Bonferroni correction, significant clusters remained for the sentence task (FNC and SPC) and the word list task (SPC only). No significant differences were detected for the white noise condition.

Overall, these findings indicate that the word list task captured the most robust and consistent group differences, particularly involving AVH+ participants. Subsequent analyses, including energy landscape modeling, were therefore focused on this task to explore neural dynamics associated with auditory verbal hallucinations.

### Functional Network Connectivity

#### Connectivity Maps: AVH+ vs CON – Word List Task

Schizophrenia with auditory verbal hallucinations (AVH) is associated with sensorimotor dysconnectivity that disrupts inner speech generation, sensory integration, and self-monitoring. In the AVH+ (N = 23) vs. control (N = 25) comparison during the word list task, functional network connectivity revealed significant clusters across the visual, sensorimotor, and subcortical networks. Within the visual network, bilateral V5/MT+, regions involved in motion perception and audiovisual integration—showed altered connectivity. Abnormal activity in these areas may reflect impaired predictive coding or excessive cross-modal binding, contributing to the externalization of hallucinated voices [34–38].

In the sensorimotor network, bilateral A4hf and A4t—subdivisions of the primary motor cortex—were implicated. A4hf, responsible for articulatory control and inner speech regulation, may show hyperactivity linked to involuntary speech generation or hypoactivity reflecting poor efference copy transmission. A4t, though less directly involved in speech, contributes to postural control and broader motor integration; its dysfunction may relate to altered bodily self-awareness in AVH [34–38].

**Fig 1.**
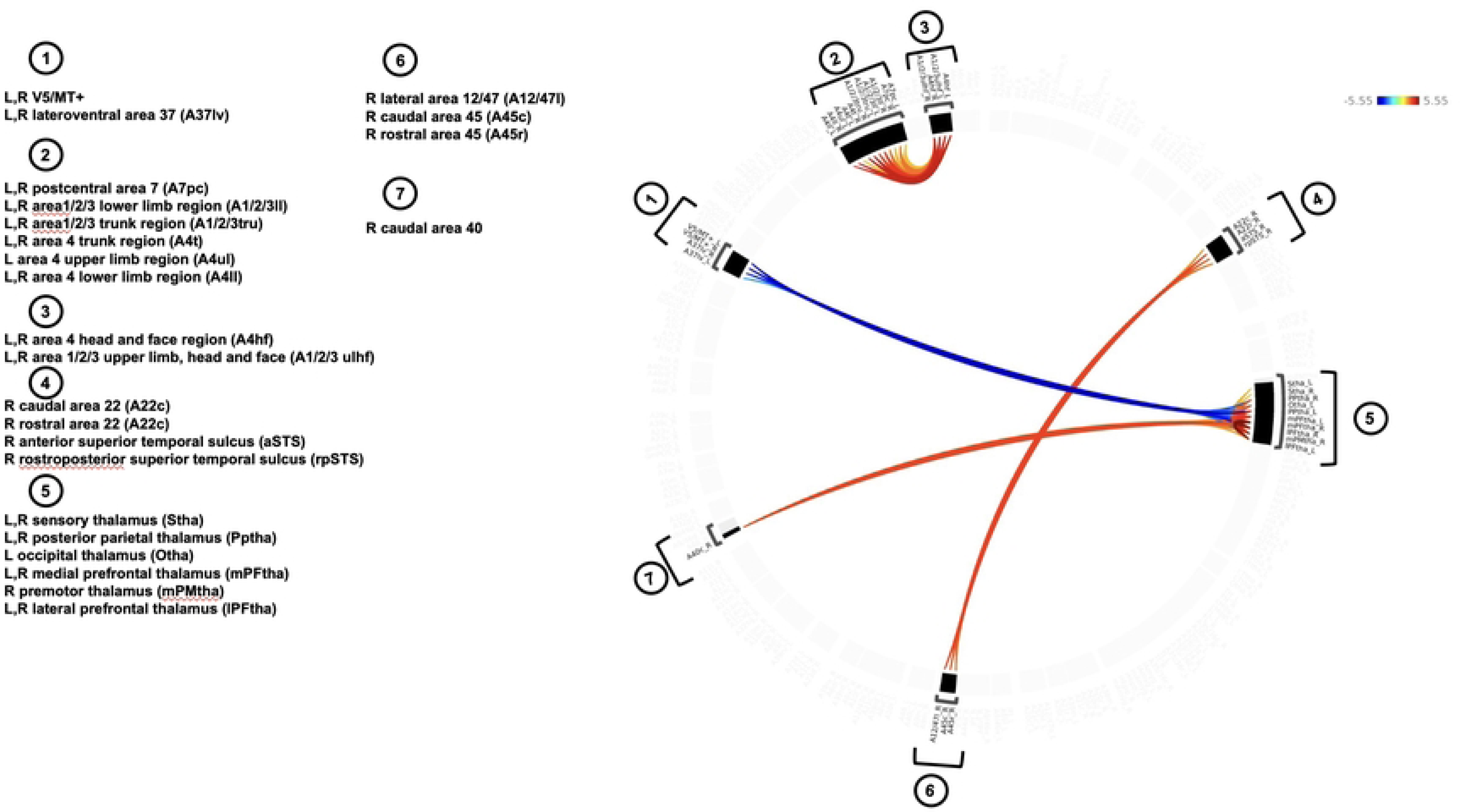
The mean functional connectivity for the between-subject [CON (*−*1) vs. AVH+ (1)] contrast presented as a connectome ring showing network of correlations for the *word lists* task: *before* Bonferroni correction computed using functional network connectivity (FNC).

**Fig 2.**
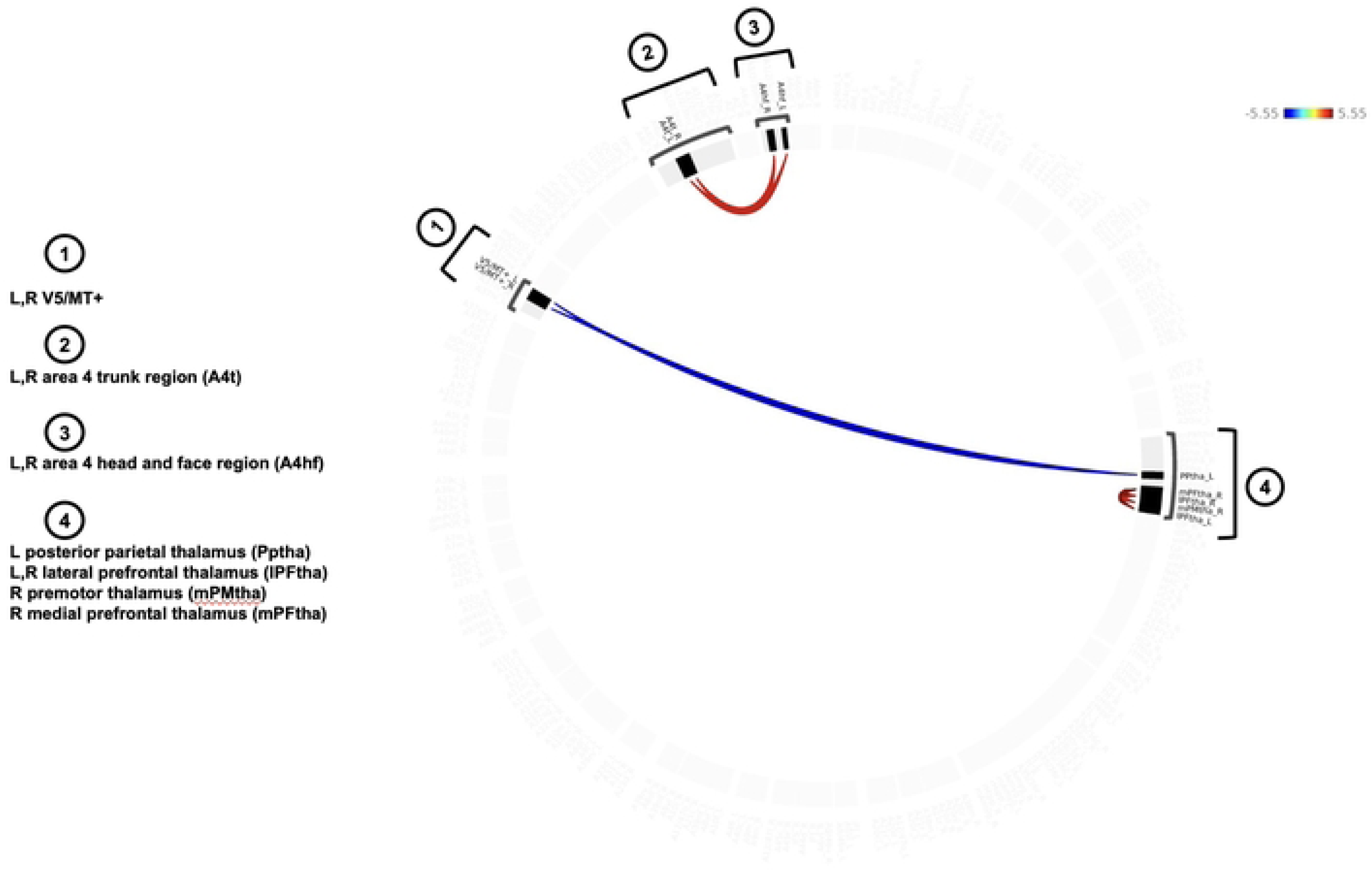
The mean functional connectivity for the between-subject [CON (*−*1) vs. AVH+ (1)] contrast presented as a connectome ring showing network of correlations for the *word lists* task: *after* Bonferroni correction, showing the reduced number of ROIs in a single brain display computed using functional network connectivity (FNC).

Subcortical findings included five thalamic ROIs. The right medial prefrontal-thalamus (mPFtha), connected to the medial prefrontal cortex, plays a role in reality monitoring and executive control. Dysconnectivity here may impair top-down regulation, leading to misattribution of internal speech. The right premotor-thalamus (mPMtha), linked to motor planning regions, may reflect disrupted sequencing and monitoring of inner speech. The left posterior parietal-thalamus (Pptha), connected to attention and sensory integration hubs, may show abnormal activity that heightens focus on hallucinated stimuli or weakens auditory filtering. Finally, the bilateral lateral prefrontal-thalamus (lPFtha), connected to the dorsolateral prefrontal cortex, is involved in cognitive control and inhibition [34–38].

### Activation Maps of Paired States: AVH+ vs Controls - Word List Task

There were 2^1^1 or 2,048 possible connectivity states among the eleven ROIs. After Bonferroni correction, four significant pairs of activation states were identified. In Figure 3, the activation maps of the paired states indicate the ROIs that were active or inactive together, and thus correlated or anti-correlated. Boxplots of the four significant pairs of activation states are shown in Figure **??**, showing that the control group has slightly lower energies than the AVH+ group, suggesting more stable connections [19, 34–38].

**Fig 3.**
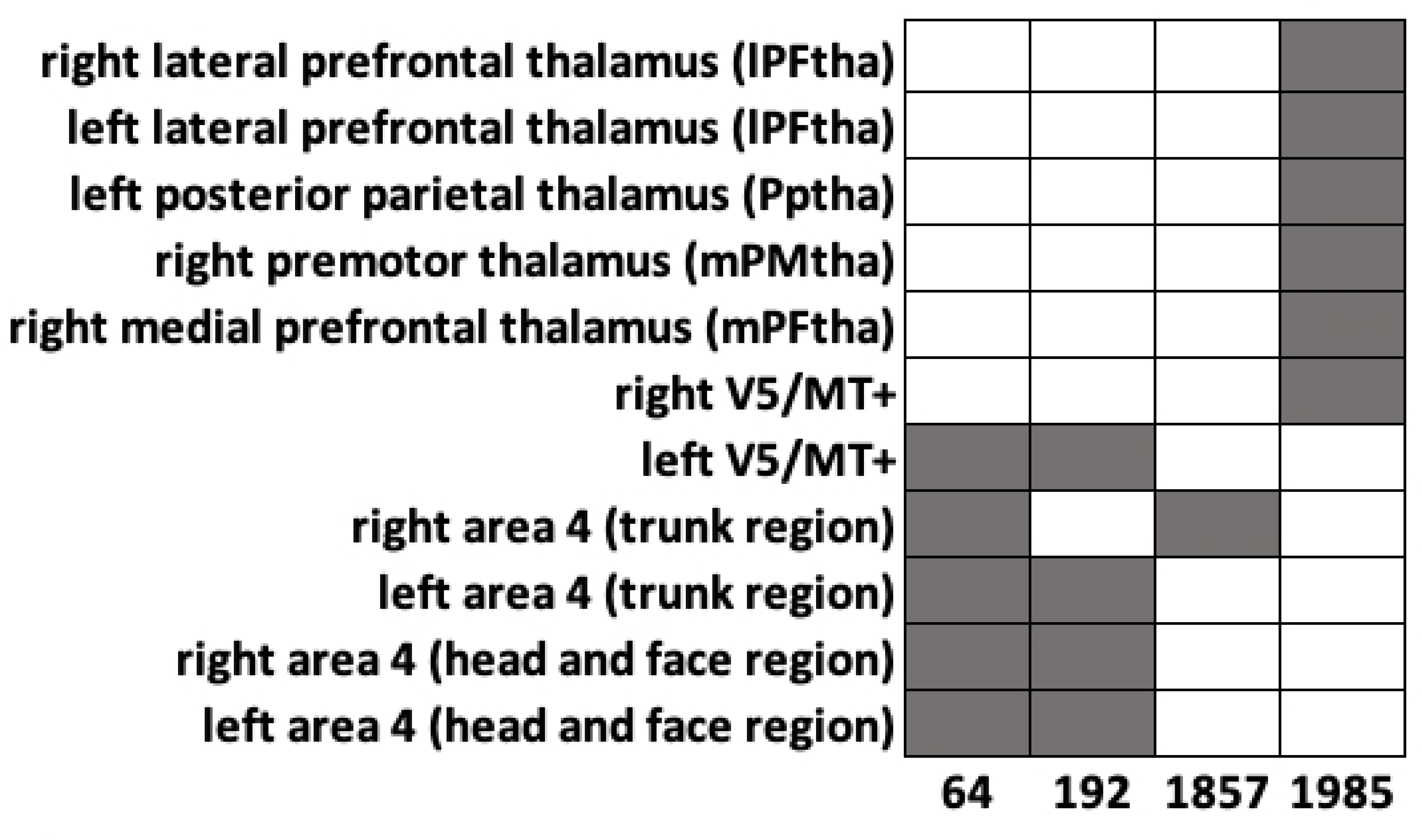
There were 2*^N^* or 2,048 possible connectivity states across the ROIs. After Bonferroni correction, four significant activation (64, 192, 1857, and 1985) states of the CON and AVH+ groups were found, with paired states 64 and 1985 also appearing as local minimums. The activation maps of the paired states indicate the ROIs that were active or inactive together, and thus correlated or anti-correlated.

**Fig 4.**
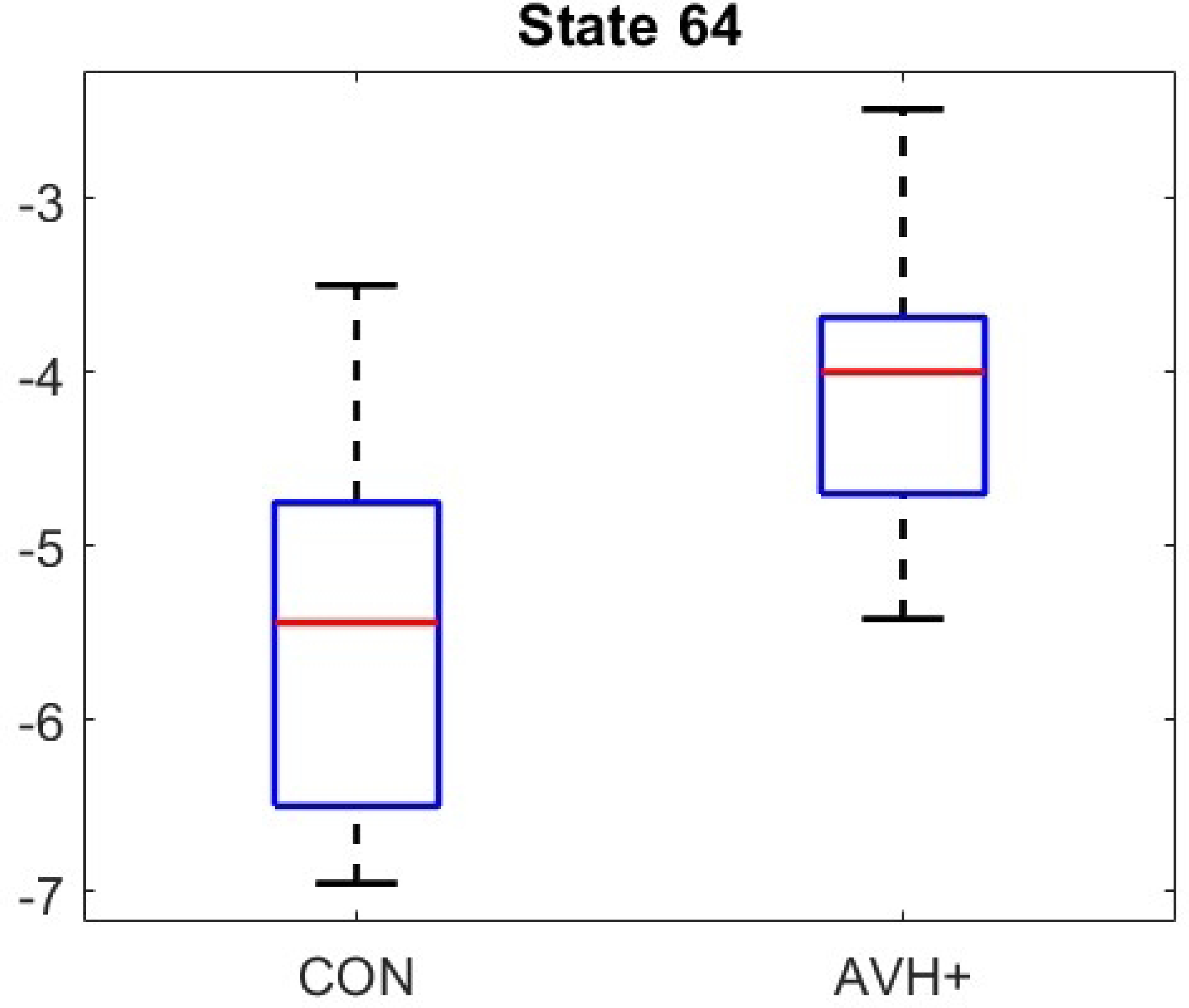
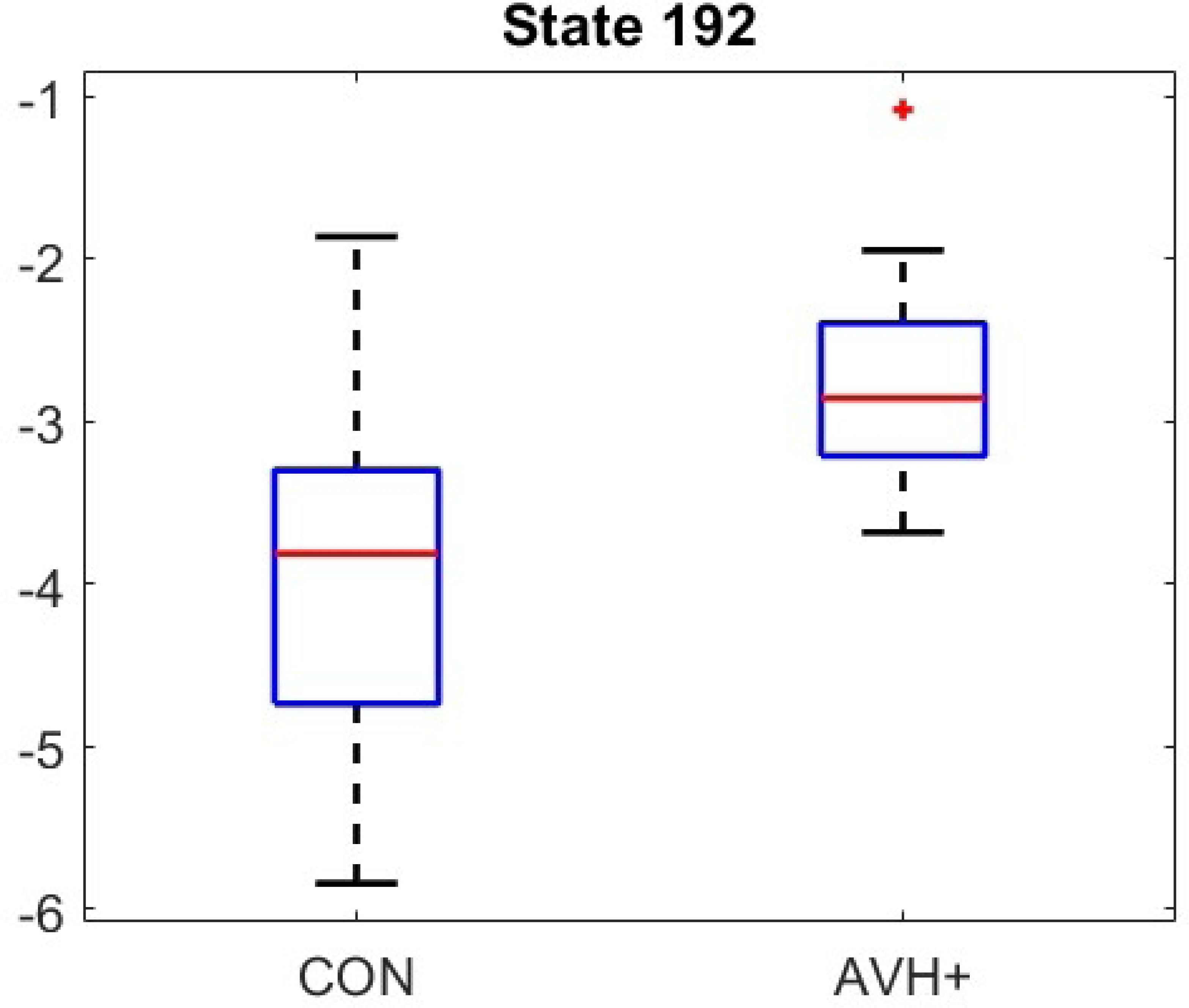
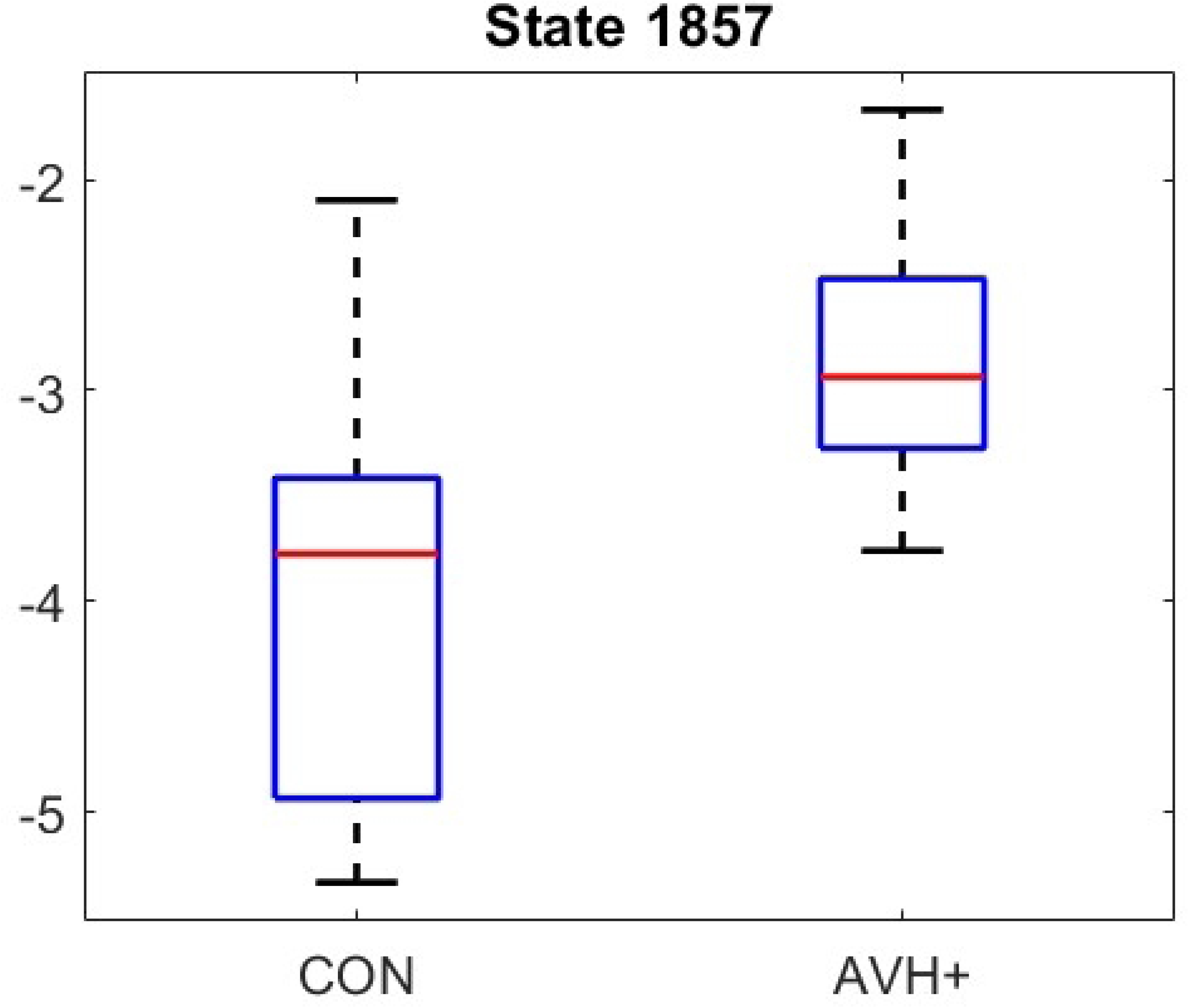
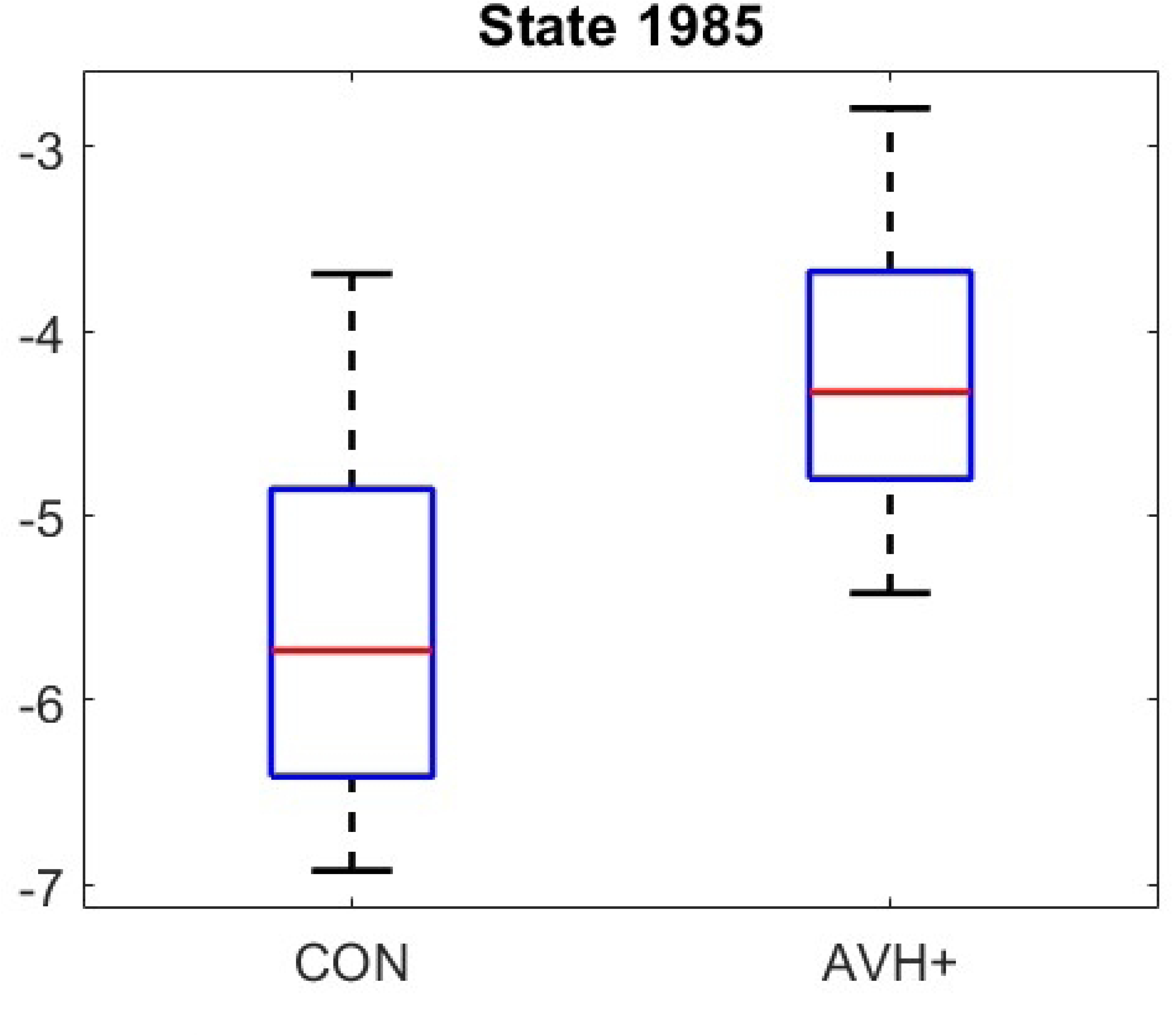
Boxplots of the mean brain energy states for the between-subject [Control (-1) vs. AVH+ (1)] contrast for the *word lists* task. Results are derived from the CONN toolbox, using the *functional network connectivity* (FNC) hierarchical clustering method and energy landscape analysis. States 64, 192, 1857, and 1985 passed the Bonferroni correction method. Paired states are complementary, stable, and potential connectivity biomarkers.

### Spatial Pairwise Connectivity

#### Connectivity Maps: AVH+ vs CON - Word List Task

Schizophrenia with AVH is associated with thalamocortical dysconnectivity, affecting sensory processing, attention, working memory, and cognitive control. In the AVH+ (N = 23) vs. control (N = 25) comparison during the word list task, spatial pattern connectivity (SPC) revealed four significant subcortical ROIs: right medial prefrontal-thalamus (mPFtha), right premotor-thalamus (mPMtha), and bilateral lateral prefrontal-thalamus (lPFtha).

The mPFtha, connected to the medial prefrontal cortex (mPFC), supports executive function and reality monitoring. Dysconnectivity in this circuit may alter top-down regulation, leading to misattribution of internal thoughts as external voices.

Hyperactivity may reflect excessive self-referential processing, while hypoactivity suggests weakened cognitive control over intrusive verbal content. The mPMtha, linked to the premotor and supplementary motor areas, plays a role in motor planning and inner speech regulation. Dysfunction here may disrupt inner speech monitoring, resulting in spontaneous internal speech without adequate sensorimotor feedback, a mechanism that contributes to AVH. The bilateral lPFtha, connected to the dorsolateral prefrontal cortex (DLPFC), is involved in working memory and inhibition of irrelevant stimuli. Dysconnectivity in this pathway may impair the suppression of internally generated speech, making it difficult for patients to distinguish self-generated thoughts from external voices [34–38].

**Fig 5.**
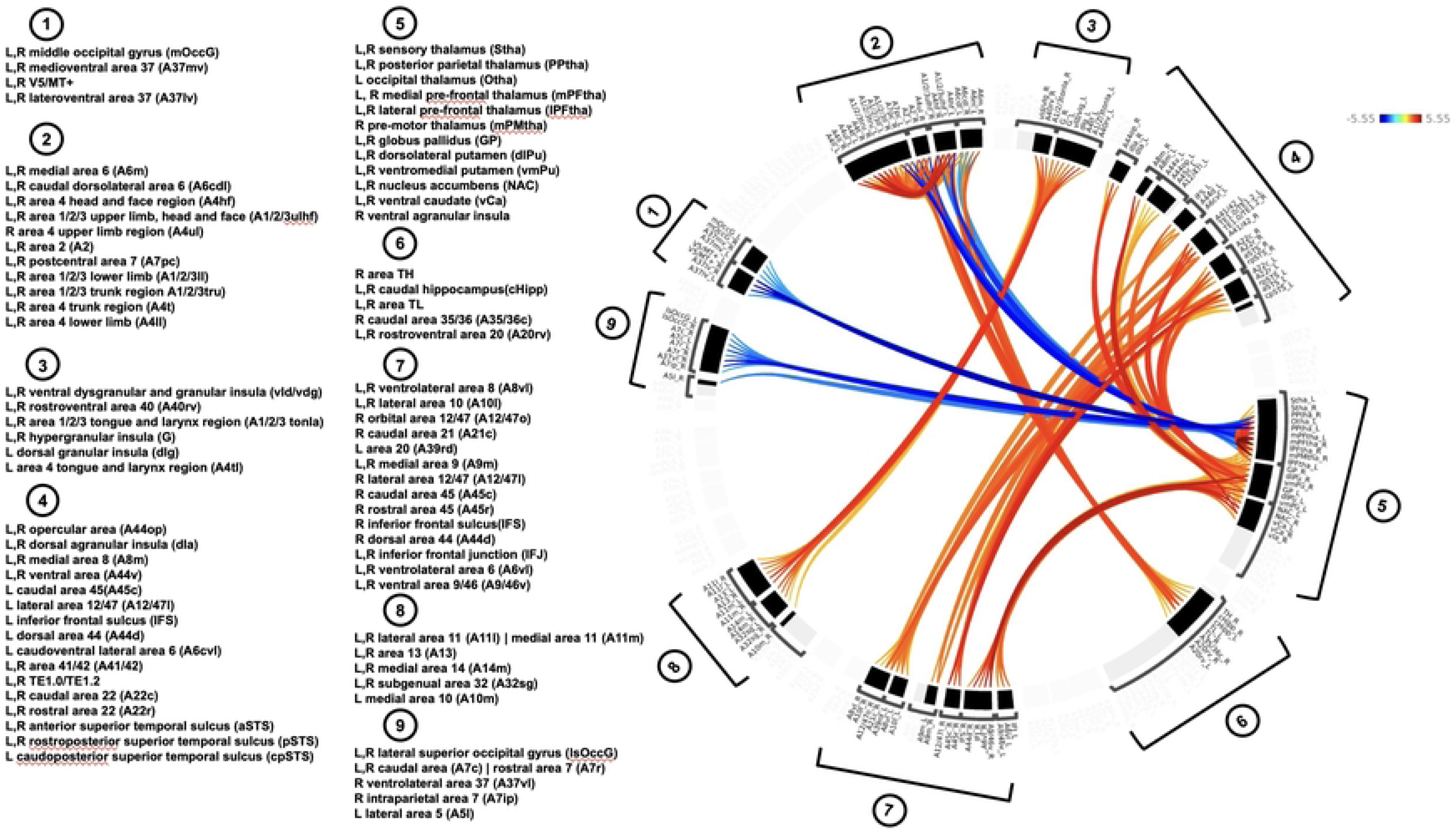
The mean functional connectivity for the between-subject [CON(*−*1) vs. AVH+ (1)] contrast presented as a connectome ring showing network of correlations for the *word lists* tasks: *before* Bonferroni correction computed using spatial pairwise connectivity (SPC).

**Fig 6.**
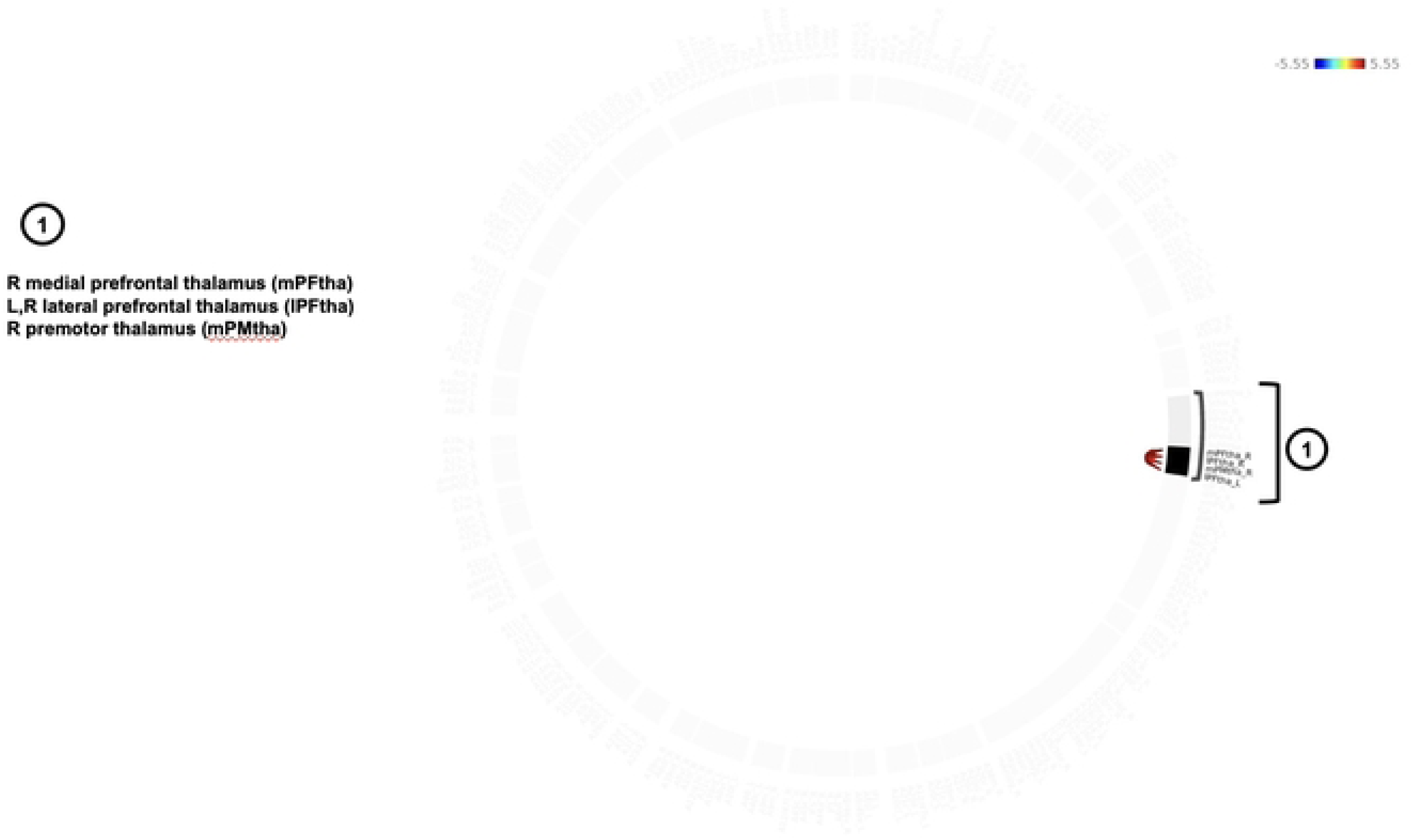
The mean functional connectivity for the between-subject [AVH- (*−*1) vs. AVH+ (1)] contrast presented as a connectome ring showing network of correlations for the *words lists* tasks: *after* Bonferroni correction, showing the reduced number of ROIs in a single brain display computed using spatial pairwise connectivity (FNC).

### Activation Maps of Paired States: AVH+ vs Controls - Word List Task

There were 2*^N^* or 16 possible connectivity states across the ROIs. After Bonferroni correction, two significant paired activation states were found. In Figure 3, the activation maps of the paired states indicate the ROIs that were active or inactive together, and thus correlated or anti-correlated. The boxplots of the two significant paired activation states are shown in Figure 8, showing that the control group has slightly lower energies than the AVH+ group, suggesting more stable connections.

### State Dynamics

State transition matrices derived from functional connectivity (FNC) and spatial pairwise connectivity (SPC) analyses revealed distinct patterns during the word list task in controls and AVH+ patients.

For FNC, of the Bonferroni-significant states 64, 192, 1857, and 1985—two states (64 and 1985) were identified as local minima. These states showed higher self-transition probabilities for controls (Table 4, than AVH + patients as shown in Table 5. Controls had longer dwell times than AVH+ patients (Table 6) and more visit counts Table 7). These states appear to represent stable, low-energy configurations associated with efficient neural processing. In contrast, AVH+ patients exhibited more frequent transitions and shorter dwell times, suggesting less stable network dynamics and elevated energy demands.

**Table 4.** State transition matrix for the Control group using functional network connectivity (FNC), for the *word lists* task.

|  | State 64 | State 192 | State 1857 | State 1985 |
| --- | --- | --- | --- | --- |
| State 64 | 0.35 | 0.06 | 0.00 | 0.00 |
| State 192 | 0.14 | 0.14 | 0.00 | 0.00 |
| State 1857 | 0.00 | 0.00 | 0.17 | 0.17 |
| State 1985 | 0.00 | 0.00 | 0.03 | 0.41 |

**Table 5.** State transition matrix for the AVH+ group using functional network connectivity (FNC), for the *word lists* task.

|  | State 64 | State 192 | State 1857 | State 1985 |
| --- | --- | --- | --- | --- |
| State 64 | 0.24 | 0.04 | 0.00 | 0.00 |
| State 192 | 0.19 | 0.06 | 0.00 | 0.00 |
| State 1857 | 0.00 | 0.00 | 0.11 | 0.05 |
| State 1985 | 0.00 | 0.00 | 0.01 | 0.26 |

**Table 6.** Comparison of average dwell time for HC and AVH+

| State | Avg. Dwell Time (HC) | Avg. Dwell Time (AVH+) |
| --- | --- | --- |
| 64 | 1.49 | 1.19 |
| 192 | 1.02 | 0.49 |
| 1857 | 0.90 | 0.74 |
| 1985 | 1.55 | 1.28 |

**Table 7.** Comparison of total visit counts for HC and AVH+

| State | Total Visit Count (HC) | Total Visit Count (AVH+) |
| --- | --- | --- |
| 64 | 253 | 82 |
| 192 | 59 | 16 |
| 1857 | 48 | 19 |
| 1985 | 301 | 82 |

SPC analysis similarly identified states 1 and 16 as local minima, with controls again showing higher self-transition probabilities (Table 8 than AVH+ patients (Table 9).

**Table 8.** State transition matrix for the HC group using spatial pairwise connectivity (SPC), for the *word lists task*.

|  | State 1 | State 16 |
| --- | --- | --- |
| State 1 | 0.64 | 0.03 |
| State 16 | 0.03 | 0.64 |

**Table 9.** State transition matrix for the AVH+ group using spatial pairwise connectivity (SPC), for the *word lists* task.

|  | State 1 | State 16 |
| --- | --- | --- |
| State 1 | 0.59 | 0.03 |
| State 16 | 0.02 | 0.59 |

Controls also had longer dwell times (Table 10) and more visit counts than AVH+ (Table 11). These patterns reinforce the idea that healthy brains tend to occupy energetically favorable and functionally efficient states for longer durations, supporting sustained attention, working memory, and reality monitoring.

**Table 10.** Comparison of average dwell time for HC and AVH+

| State | Avg. Dwell Time (HC) | Avg. Dwell Time (AVH+) |
| --- | --- | --- |
| 1 | 2.81 | 2.44 |
| 16 | 2.76 | 2.45 |

**Table 11.** Comparison of total visit counts for HC and AVH+

| State | Total Visit Count (HC) | Total Visit Count (AVH+) |
| --- | --- | --- |
| 1 | 2156 | 1697 |
| 16 | 2163 | 1746 |

Physiologically, lower energy states may reflect more metabolically efficient configurations, while longer dwell times suggest greater neural stability and resistance to perturbation. These dynamics are consistent with robust cognitive control and the suppression of intrusive verbal content—functions often impaired in schizophrenia.

The instability and elevated energy observed in AVH+ patients may reflect a breakdown in predictive coding, where top-down expectations override bottom-up sensory input, resulting in misattributed internal speech. Modeling these brain state transitions and energy landscapes could inform biologically grounded AI architectures capable of filtering internally generated noise and maintaining representational stability.

Together, these results highlight distinct physiological and computational differences in brain state stability and network efficiency between patients and controls. Future work could explore how these states relate to behavioral performance and whether they can be modulated to improve cognitive function or reduce hallucinatory symptoms.

**Fig 7.**
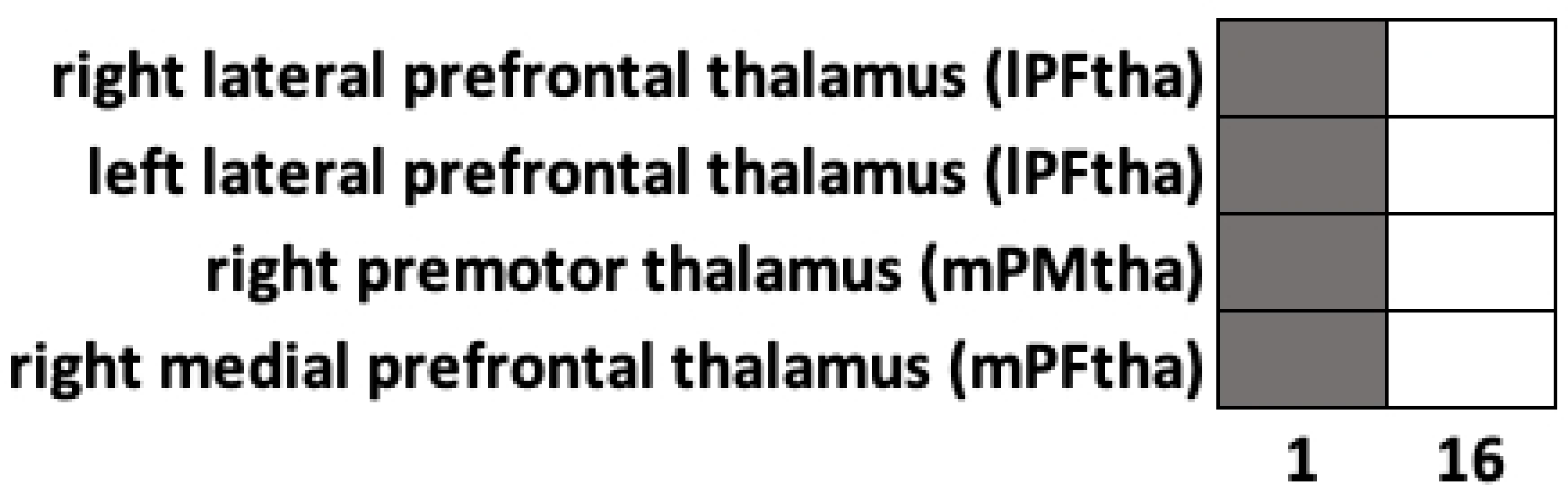
There were 2*^N^* or 16 possible connectivity states across the ROIs. After Bonferroni correction, two significant activation states of the CON and AVH+ groups, were identified as 1 and 16, with paired states 1 and 16 also appearing as local minimums. The activation maps of the paired states indicate the ROIs that were active or inactive together, and thus correlated or anti-correlated.

**Fig 8.**
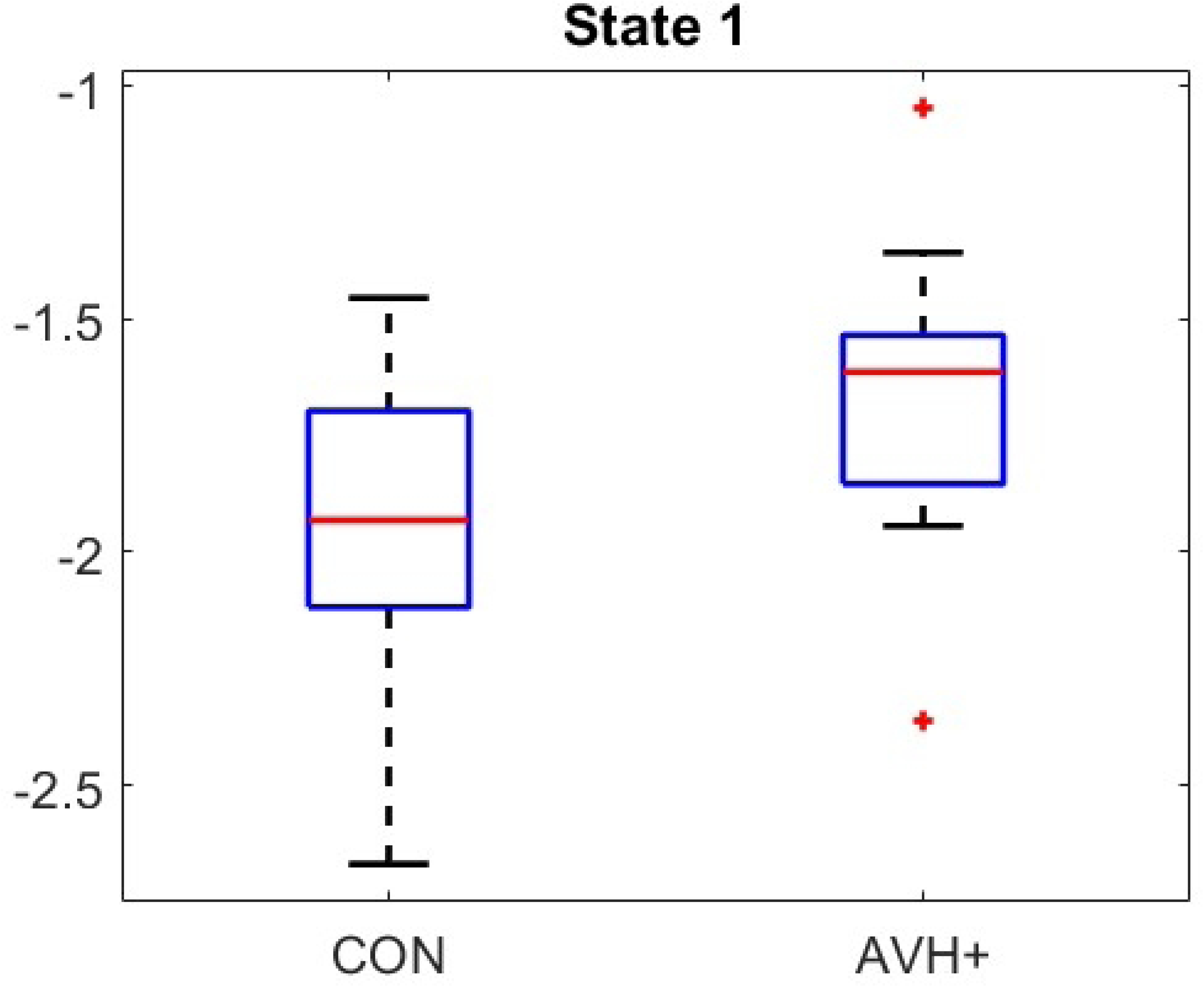
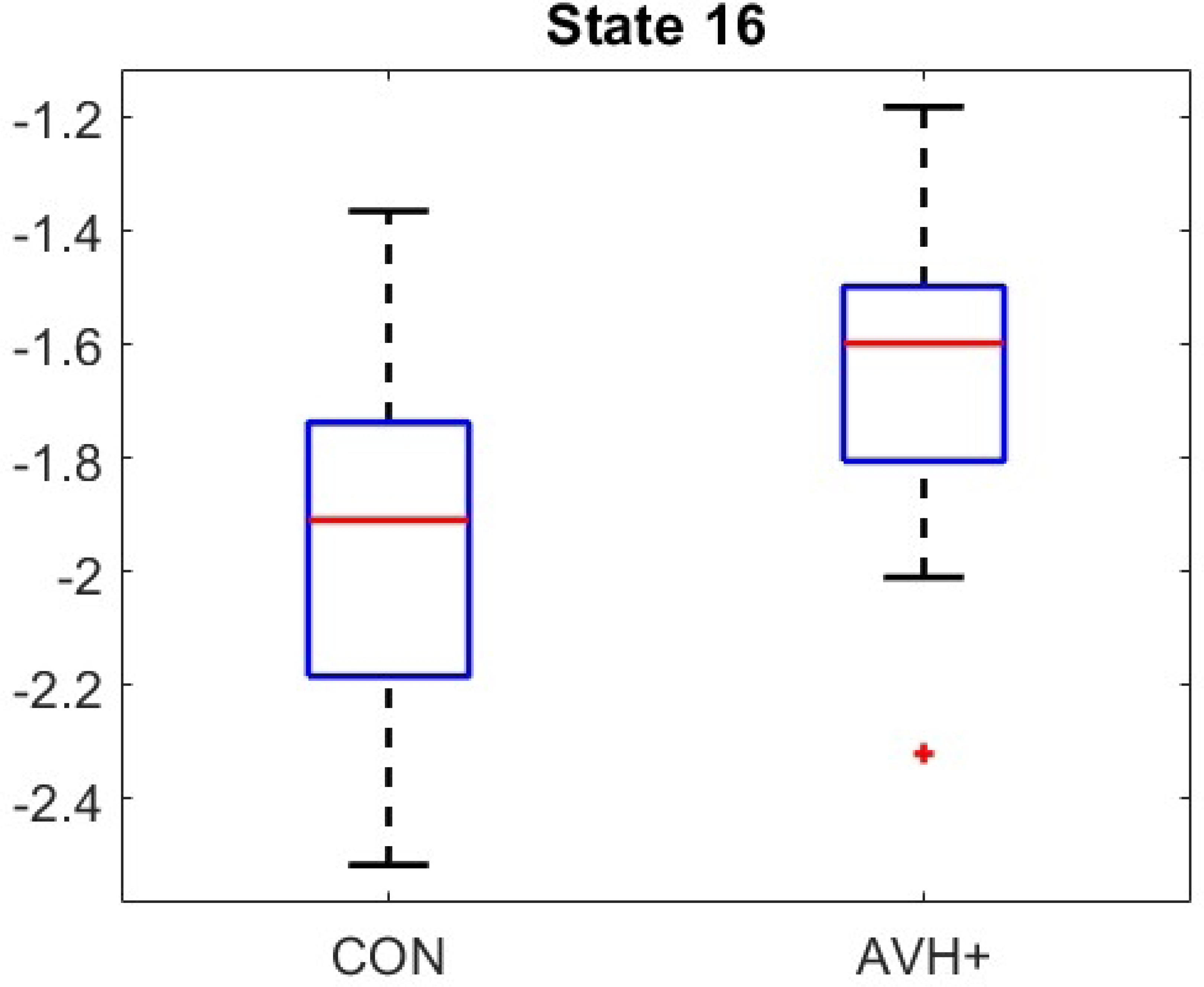
Boxplots of the mean brain energy states for the between-subject [Control (-1) vs. AVH+ (1)] contrast for the *word lists* task. Results are derived from the CONN toolbox, using the *spatial pairwise connectivity* (SPC) hierarchical clustering method and energy landscape analysis. Two pairs of states passed the Bonferroni correction method, and states 1 and 16 are also local minimums. Paired states are complementary, stable, and potential connectivity biomarkers.

## Discussion

### FNC Physiology: CON vs. AVH+ – Word Lists Task

This study examined sensorimotor and thalamocortical dysconnectivity in schizophrenia patients with auditory verbal hallucinations (AVH), focusing on how disrupted brain networks contribute to inner speech generation, motor perception, and reality monitoring. Functional network connectivity (FNC) analysis during the word list task revealed significant group differences between AVH+ patients (N = 23) and healthy controls (N = 25), identifying key regions across the visual, sensorimotor, and subcortical networks

Functional Network Connectivity (FNC) analyses examined sensorimotor and thalamocortical dysconnectivity in schizophrenia patients with auditory verbal hallucinations (AVH), focusing on how disrupted brain networks contribute to inner speech generation, motor perception, and reality monitoring. Altered connectivity in these systems has been widely implicated in AVH pathophysiology, particularly through failures of predictive coding, corollary discharge, and self-monitoring mechanisms [11–13, 39]. Functional network connectivity (FNC) analysis during the word list task revealed significant group differences between AVH+ patients (N = 23) and healthy controls (N = 25), identifying key regions across the visual, sensorimotor, and subcortical networks.

### Visual Network Dysfunction

The left and right V5/MT+ regions—critical for motion processing and audiovisual integration—showed altered connectivity in AVH+ patients. Abnormal MT+ responses have been observed in schizophrenia and are associated with impaired motion perception and sensory integration [40]. MT+ interacts with V1, STS, parietal, and prefrontal regions, supporting predictive coding and multimodal binding [38]. Dysfunction here may reflect excessive audiovisual coupling or weakened sensory prediction, both of which can enhance the externalization of internally generated auditory content. Abnormal cross-modal integration has also been described in schizophrenia, further supporting this interpretation [41–44].

### Sensorimotor Network Dysconnectivity

ROIs in the primary motor cortex (M1)—specifically the head/face (A4hf) and trunk (A4t) regions—were implicated in AVH+ patients. A4hf contributes to articulatory control and subvocal speech production. Previous work links abnormal motor cortex activation and involuntary subvocalization to AVH [45], consistent with theories of defective copy signaling [39]. Hyperactivity may amplify covert speech, while hypoactivity may weaken motor predictions, impairing the recognition of self-generated inner speech. A4t, involved in postural and bodily integration, may reflect broader embodied disturbances sometimes observed in schizophrenia.

### Subcortical Network Disruption

Thalamic ROIs—including the right medial prefrontal – thalamus (mPFtha), right premotor – thalamus (mPMtha), left posterior parietal – thalamus (Pptha), and bilateral lateral prefrontal – thalamus (lPFtha)—support executive function, motor planning, and sensory integration.

Extensive evidence shows that thalamocortical circuits are altered in schizophrenia [35–37]. Disruption in mPFtha and mPMtha may impair regulation of internal speech and cognitive control, while dysfunction in Pptha and lPFtha–connected to posterior parietal cortex and DLPFC –may weaken attentional gating and working memory functions [34, 35]. Together, these abnormalities compromise the efference copy mechanism, leading to misattribution of internal speech as external [12, 39].

### State-Based Biomarkers and Energy Dynamics

Energy landscape analysis identified several Bonferroni-significant activation states (64, 192, 1857, 1985), that differentiated AVH+ from controls. Similar approaches have previously been shown to capture unstable attractor dynamics in psychiatric populations [14, 15, 19, 31–33].

Controls exhibited lower energy and longer dwell times, indicating more stable network configurations, whereas AVH+ patients showed higher energy and reduced dwell time—patterns consistent with neural instability and impaired cognitive filtering.

### SPC Physiology: Control vs. AVH+ – Word Lists Task

Spatial pairwise connectivity (SPC) analyses further highlighted thalamocortical disruptions. Four subcortical ROIs—mPFtha, mPMtha, and bilateral lPFtha—showed significant differences between AVH+ and controls.

### Medial Prefrontal-Thalamus (mPFtha)

mPFtha connects with the medial prefrontal cortex, supporting executive function, salience processing, and reality monitoring. Thalamic–prefrontal dysconnectivity is a well-established feature of schizophrenia and is associated with cognitive control and self-monitoring deficits [34–37]. Altered mPFtha activity may thus impair the suppression of intrusive internal speech, contributing to AVH.

### Premotor-Thalamus (mPMtha)

This region supports motor planning and inner speech regulation. Abnormal premotor–thalamic signaling can disrupt the internal monitoring of self-generated speech, paralleling findings of reduced corollary discharge in AVH [39, 45].

### Lateral Prefrontal-Thalamus (lPFtha)

Connected to the DLPFC, lPFtha mediates working memory and inhibitory control. Schizophrenia is characterized by reduced DLPFC engagement and attentional dysregulation [34, 35]. Dysconnectivity in this pathway may impair the filtering of internally generated verbal content, reinforcing the externalization of inner speech.

### State-Based Biomarkers and Energy Dynamics

SPC revealed four Bonferroni-significant activation states (1, 4, 13, 16), with states 1 and 16 serving as local minima—potential biomarkers of network stability. Controls exhibited lower energy and longer dwell times, consistent with efficient thalamic–prefrontal dynamics. In contrast, AVH+ patients showed elevated energy and unstable transitions, akin to attractor dynamics in computational systems when internal representations become noisy and unreliable [30–33].

### Interpretation and Integration

Together, the FNC and SPC analyses converge on a central conclusion: AVH arise from disrupted thalamic–sensorimotor–prefrontal circuitry. These circuits support predictive coding, efference copy, and reality monitoring—mechanisms crucial for distinguishing self-generated thoughts from external stimuli [11–13, 38, 39]. The instability observed in energy landscape dynamics aligns with theories of dysregulated attractor states in schizophrenia, where noisy internal predictions produce perceptual distortions resembling hallucinations [14, 15, 19, 31–33]. These findings highlight network-level biomarkers of AVH and underscore the promise of task-based connectivity and energy-based modeling for characterizing perceptual disturbances in schizophrenia.

## Conclusion

This study provides converging evidence that auditory verbal hallucinations (AVH) in schizophrenia arise from widespread dysconnectivity across visual, sensorimotor, and thalamocortical networks. Task-based fMRI revealed that AVH+ patients exhibit unstable brain state transitions, elevated energy dynamics, and reduced dwell times—hallmarks of neural instability and impaired cognitive filtering. Functional network connectivity (FNC) and spatial pairwise connectivity (SPC) analyses identified key disruptions in regions responsible for inner speech generation, motor feedback, and executive control, including V5/MT+, A4hf, mPFtha, mPMtha, and lPFtha.

These findings support a systems-level account of hallucinations as emergent phenomena driven by dysregulated predictive coding and impaired reality monitoring. The failure of the efference copy mechanism, combined with hyperactive sensory integration and weakened top-down inhibition, renders internally generated speech indistinguishable from external auditory input. This breakdown in generative control mirrors attractor instability in artificial neural networks, where noisy internal representations can produce fluent but spurious outputs—akin to hallucinations in both biological and computational systems.

Future research is expected to delve deeper into the neural mechanisms underlying predictive coding and generative control, potentially identifying specific circuit-level codes that govern cognitive filtering and sensory integration. By mapping how neural information processing might resemble computational principles—such as those seen in advanced artificial systems—this line of inquiry may bridge gaps between neuroscience and machine learning.

Such interdisciplinary efforts hold promise not only for advancing our understanding of schizophrenia and other neuropsychiatric disorders but also for informing the development of novel biomarkers and targeted therapies. Ultimately, exploring these predictive architectures at both biological and computational levels could pave the way for innovative interventions and inspire new paradigms in artificial intelligence design, grounded in the complexities of brain function.

## Data Availability

The data underlying the results presented in this study are publicly available from the OpenNeuro repository: https://openneuro.org/datasets/ds004302/versions/1.0.0/metadata. All analyses reported in this manuscript were conducted using these existing datasets.

## Acknowledgments

We thank our coauthors for their support, feedback, and collaboration throughout the development of this work. We also extend our appreciation to colleagues whose discussions and insights contributed to the refinement of the study. We thank Soler-Vidal et al. and their collaborators for making the dataset used in this research publicly available, and we acknowledge the original participants and research staff involved in data collection and study coordination.

## Notes

### Competing Interest Statement

The authors have declared no competing interest.

### Author Declarations

This study used secondary data obtained from publicly available sources. No new human subjects were directly involved, and the original data collection was conducted under appropriate ethical oversight

